# Comparison of Japanese nurse practitioner-led care and physician trainees-led care on patients’ length of stay in a secondary emergency department: A retrospective study

**DOI:** 10.1101/2020.10.26.20215897

**Authors:** Keiichi Uranaka, Hitoshi Takaira, Ryoji Shinohara, Zentaro Yamagata

**Author notes:** Corresponding author (KU). These authors contributed equally to this work (Hitoshi Takaira, Ryoji Shinohara, Zentaro Yamagata).

## Abstract

**Objectives:** We compared nurse practitioner-led care and physician trainees-led care on patients’ length of stay in a secondary emergency department in Japan.

**Methods:** This observational research utilized a secondary data analysis of medical records. Participants (N = 1,419; mean age = 63.9 ± 23.4 years; 52.3% men) were patients who were transferred to the emergency department by an ambulance between April 2016 and March 2018 in western Tokyo. Multiple linear regression analyses were performed with the length of stay as the dependent variable and the factors related to the length of stay, including medical care leaders, as the independent variable to compare Japanese nurse practitioner-led care and physician trainees-led care on patients’ length of stay.

**Results:** Approximately half of the patients (n = 763; 53.8%) received Japanese nurse practitioner-led care. Patients’ length of stay was significantly shorter by six minutes in the Japanese nurse practitioner-led care group than the physician trainees-led care group.

**Conclusion:** Patients’ length of stay was significantly shorter by six minutes in the Japanese nurse practitioner group than the physician trainees’ group. This time difference suggests that the medical care led by Japanese nurse practitioners is more efficient. In the future, the cost-effectiveness of Japanese nurse practitioner medical care, safety, and patient satisfaction should be examined in a multi-institutional joint study.

## Introduction

Demand for emergency medicine is increasing annually, with an increase in the number of presentations to hospitals [1]. However, owing to the uneven distribution of physicians and the enforcement of work training time limits for trainee physicians [2–4], the workload of emergency physicians is high. In addition, owing to the serious shortage of emergency department staff, emergency departments (EDs) face problems such as increased waiting time and length of stay for patients. The extension of the length of stay is associated with higher mortality rates [5–7] and prolonged hospital length of stay [8], which increases patients’ medical costs and reduces patient satisfaction [9–10].

To solve these problems, health services have been remodeled significantly in response to healthcare demands. Increased public uptake of care provided by specialists, including nurse practitioners (NPs), can reduce care latency [11]. In primary care and emergency medicine, NPs are being introduced worldwide as a solution to these noted problems [12–14], and patients’ waiting time and dissatisfaction are declining [15].

NPs are registered and certified nurses who have been educated to function autonomously and cooperatively in advanced and extended clinical roles. The Advanced Practice Nursing Regulations Consensus Model in 2008 provides primary, continuous, and comprehensive care— including comprehensive medical history; physical examinations and other health assessments and screening activities; the diagnosis, care, and management of acute and chronic patients and diseases; order and conduct laboratory examinations and imaging examinations; interpret examination results; prescribe drugs; use medical devices; and provide explanations to patients [16]. The International Council of Nurses [17] also defines an NP as a registered nurse with expert knowledge, complex decision-making skills, and clinical competence—with legislated extensions for expanded practice. NPs have acquired advanced knowledge and skills regarding pathophysiology, physical assessment, and pharmacology, which undergraduate programs in many countries cover to some degree, but not in-depth as in the NP programs. They are clinically trained, hold master’s degrees, and are certified. The number of NPs is increasing annually: there are 290,000 in the United States [18], 5,697 in Canada [19], and 1,839 in Australia [20]. In addition, emergency care research has revealed that NPs provide a valuable, safe, and effective service [21–23].

When patients receive care from NPs and physicians in EDs, their length of stay is shorter than those who receive care from physicians only [12–14]. The claim in these studies was that NPs had a beneficial effect on patients’ length of stay in the ED. However, other previous studies revealed no significant difference in patients’ length of stay in the ED between an NP-provided care group and a physician-provided care group [12,24]. Others demonstrated that the medical hours of an emergency physician group were significantly shorter than that of an NP group [25,26]. As such, there is no unified view on the relationship between NP-provided care and patients’ length of stay in EDs.

In Japan, emergency medicine faces new social issues owing to the rapidly aging population [27] and the increasing medical demand as in many countries. As a solution, Japanese nurse practitioners (JNPs) are assigned to the field of emergency care—often leading the medical care and sharing and shifting tasks with physicians. JNPs commenced education in 2008 in a postgraduate master’s degree program [28]. JNP students will intensively study advanced medical and nursing education and acquire the ability to assist in a broader scope of practice than nurses. Currently, 11 postgraduate schools are members of the Japanese Organization of Nurse Practitioner Faculties, and the students must pass the certification qualification test to be certified as a JNP. By March 2020, 487 nurses have been certified by the Japanese Organization of Nurse Practitioner Faculties. However, in Japan, no formal legislative status has been established for NPs [29], and JNP certification is not a national qualification. The scope of practice for JNPs is similar to that of US NPs in restricted practice. Currently, JNP must not prescribe and practice medicine without a physician’s instructions. There are two types of physician’s instructions: specific instructions and comprehensive instructions. Nurses assist in medical action under the physician’s specific instructions. They must not take medical actions that carry the risk of harming a person’s health if it is not done by a physician. On the other hand, JNPs can perform specific actions (e.g., arterial punctures, correction of dehydration by intravenous drip, and adjusting the respiratory settings) based on comprehensive instructions from physicians and procedures prepared by physicians.

In EDs, JNPs have acquired and practiced the knowledge, skills, and thought processes necessary for medical care, such as physical examinations, clinical reasoning, differential diagnosis, test selections, and the interpretation of test results. However, few previous studies have examined the care outcomes of JNPs’ care in the EDs. Although the scope of practice of JNPs is not yet at an international standard, there is a need to generate evidence for the development of JNPs under current law.

A reduction of the length of stay in the EDs may contribute to higher patient satisfaction [9-10], lower mortality rates [5-7], and shorter costs [9-10] and hospital length of stay [8]. Therefore, we evaluated the efficiency of JNP’s care in a Japanese ED using an objective measure. The objective of this study was to compare JNP-led care and physician trainees-led care on patients’ length of stay in a secondary ED.

## Methods

### Design

This was an observational study with a secondary data analysis of medical records.

### Setting

This study was conducted in a medical institution in western Tokyo. At the target facility, the ED was available 24 hours a day to accept emergency patients. The JNP care model at the target facility is a collaborative model focusing on managing patients in a secondary ambulance service. During day shifts, both a JNP and physician trainees involved in care under the instruction and supervision of emergency physicians share patient information closely with emergency physicians. Regarding the assignment of patients, it is determined based not on the patients’ disease or condition but on the rotation system of the medical team in the ED. There is no waiting time and no triage because the patient arrived and the JNP or physician trainees-led care of the patient began. Two nurses who were residents and in charge of the EDs worked with JNP and physician trainees during the day shift.

### Participants

Key inclusion criteria were patients who were transferred to the secondary ED by an ambulance during the day shift (8:30 to 17:15) between April 2016 and March 2018. The reason for this criterion was that the ED at the target facility only accepted patients transported by ambulance. In addition, because the JNP worked only the day shift, we limited patients transported on the day shift to control for confounding due to differences in the work shift in which they were transported and excluded patients who were transferred on weekends and holidays.

### Data collection

The date of this study ware collected from the medical records. The records described by JNPs and physician trainees are officially recorded in patients’ electronic medical records with the approval of the emergency physicians. Electronic medical records are a platform used by medical professionals as well as administrative and accounting staff. We extracted the data of patients’ characteristics: their admission time to the ED; the presence of trauma, testing, and medical care; the discharge time from the ED; the number of tertiary emergency patients (TEPs) at the time of the participants’ arrival; and the consultations with other departments. These data were entered into a research dataset. In addition, the data used in the analysis were verified among the co-authors for transcription errors from the medical records to ensure reliability.

### Measurements

#### Length of stay

The length of stay was the primary outcome in this study. The length of stay in the ED was calculated as the difference between the time the patients were admitted and discharged to the ED in minutes. The admission time of patients was recorded by the ambulance team and entered as the reception time in the patients’ medical records. Patients’ discharge time was recorded in the medical record by the emergency nurses.

#### Care leaders

Those who recorded the following initial practices were deemed to have led the care; clinical reasoning, differential diagnosis, test selection, interpretation of test results, tentative diagnosis, and decision on a course of treatment in the ED. The JNP had a rigorous checklist of patients assigned to initial care for annual activity reports to the hospital. We classified care leaders based on the checklist and reviewed the medical records with the researcher and JNP to verify the classification. As it is often urgent to treat a patient in an ED, the JNP may lead the care, and a physician trainee records it, and vice versa. Therefore, it does not matter who wrote the medical record.

The JNP had 11 years of experience as a nurse and three years of experience as a JNP after completing the Japanese Organization of Nurse Practitioner Faculties -accredited postgraduate master’s degree and was dedicated to the ED for the period under study. The physician trainees were licensed to practice medicine as a physician for less than two years and in charge of the ED during the rotation period. If the EDs were too crowded, physician trainees were in charge of the outpatients in parallel with the JNP. When a JNP was off duty or absent, physician trainees played the role of care on behalf of the JNP in the ED. The work and thinking processes of JNPs and physician trainees were almost identical (Table 1).

**Table 1.** Work and thinking process flow of Japanese nurse practitioner and physician trainees in the emergency department

| Flow | Japanese nurse practitioner | Physician trainees |
| --- | --- | --- |
| Entering emergency department | Interview, physical examination, vital signs measurement |  |
|  | Clinical reasoning |  |
|  | Differential diseases |  |
|  | Selection of necessary tests, Explanation of tests |  |
|  | Test order (approval by emergency physician) |  |
|  | Interpretation of test results |  |
|  | [JNP] Provisional diagnosis / [Physician trainees] Diagnosis |  |
|  | Treatment and Medication (approval by emergency physician) |  |
|  | Consults with other departments |  |
| Leaving emergency department | Decision to admit, Coordination of hospitalization |  |
|  | Under the physician's instruction | Under the physician's supervision |

#### Other factors related to the length of stay

For factors related to the length of stay, based on earlier research, we examined care leader type [30,31]; patients’ age and sex [31,32]; whether they received clinical tests including x-ray photographs (XPs), computed tomography (CT), magnetic resonance imaging (MRI) [31], and blood tests [33]; catheterization [34], endoscopy, and trauma [32] characteristics; outcomes (i.e., return home or hospitalization, including transfers); the number of consultations with other departments; and the number of tertiary emergency patients when the participants arrived at the hospital. The number of tertiary emergency participants may affect the length of stay of secondary emergency patients [35]. Tertiary emergency patients require hospitalization and human resources such as physicians and nurses, and they are given priority over secondary emergency patients for testing. Therefore, secondary emergency patients often wait longer for tests than tertiary emergency patients.

### Analysis

We calculated descriptive statistics, including a simple tabulation for each variable and mean (± standard deviation) of the length of stay. The correlations between the variables were calculated using Spearman’s correlation coefficients, the tetrachoric correlation coefficient [36], and the point-biserial correlation coefficient [37] depending on the type of scale. In addition, simple linear regression analyses were performed with the length of stay as the dependent variable and the factors related to the length of stay as the independent variable. Then, a multiple linear regression analysis was performed. P-values < .05 were deemed significant. Stata version 15 (Stata Corp LLC, Lakeway, TX, USA) was used for all statistical analyses.

### Ethics Considerations

This study received ethical approval from the Institutional Review Board of the hospital (no. 2018-24) and the university (no. 2072). Informed consent was obtained in the form of opt-out on the hospital’s website. No participants refused to participate.

## Results

### Participants’ characteristics

The number of participants analyzed was 1,419, and the patients’ length of stay ranged from 6 to 486 minutes (mean = 142.9, SD = 68.6 minutes; median = 134, interquartile range = 88 minutes).

The patients’ mean age was 63.9 (SD = 23.4) years. 52.3% were men, and 47.7% were women. Further, 53.8% received JNP care, and 46.2% did not, 62.9% of the participants were non-traumatic, and 37.1% were traumatic, and 1,052 (74.1%) had no TEPs when they arrived at the hospital. The outcomes were 50.0% for both returning home and hospitalization, including transfer to another hospital. There were 340 patients (24.0%) who did not see other departments, and most (88.6%) had blood tests. In contrast, 0.2% had an endoscopy, 8.9% received an MRI test, and 0.4% received a catheterization. There was significant differences between the JNP and physician trainees-led care groups in consultation (p<.001) and blood tests (p=.004) (Table 2).

**Table 2.**
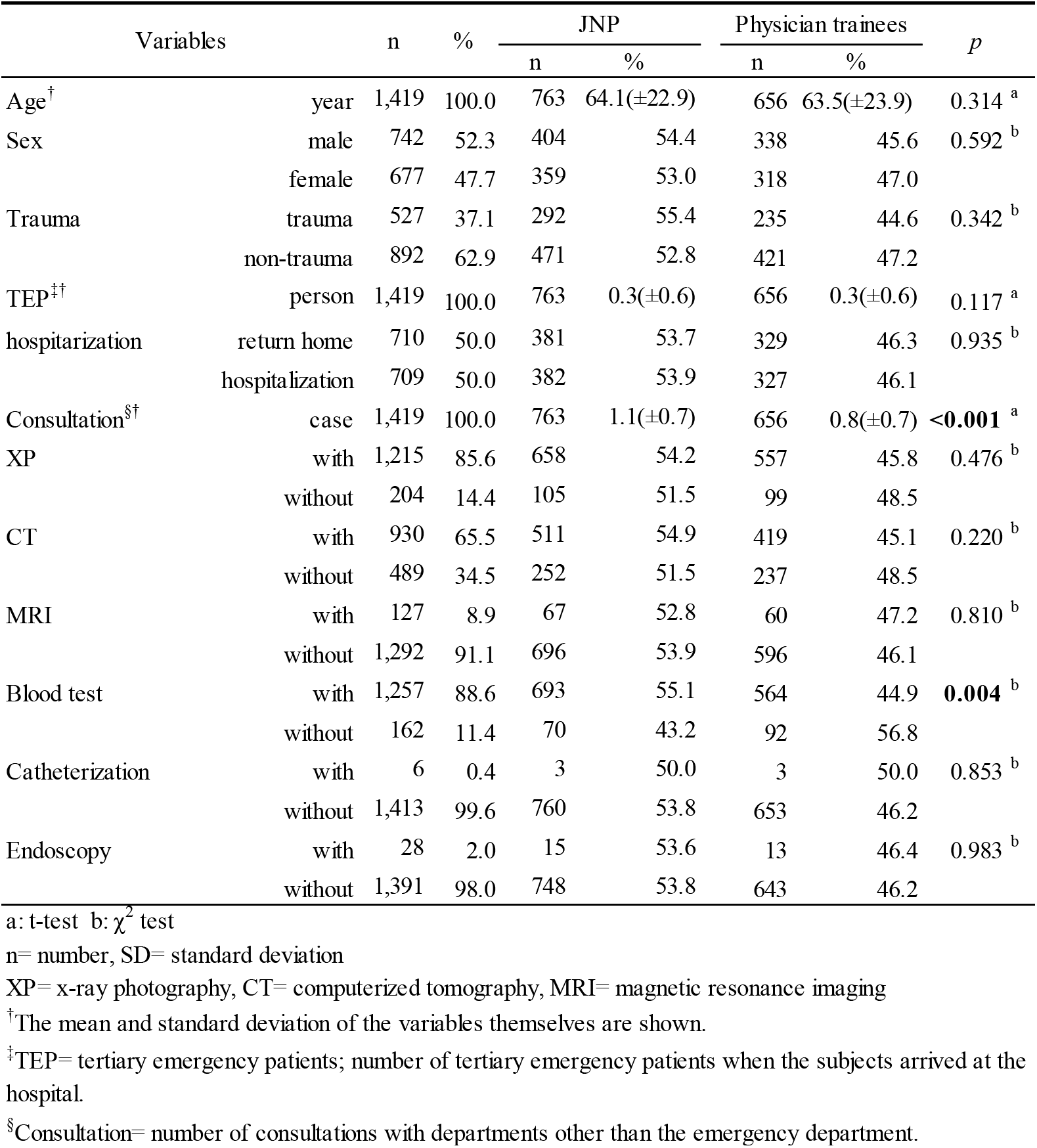
Distribution of participant characteristics by care leader (n=1 419)

There was no significant difference in the comparison of LOS in JNP-led care group and physician trainees-led care group by participant characteristics (Table 3).

**Table 3.** Comparison of LOS in JNP-led care group and Physician trainees-led care group by participant characteristics (n=1 419)

| Variables | n | % | JNP |  |  | Physician trainees |  |  | p |
| --- | --- | --- | --- | --- | --- | --- | --- | --- | --- |
|  |  |  | n | Mean | SD | n | Mean | SD |  |
| total | 1,419 | 100.0 | 763 | 143.3 | 69.9 | 656 | 142.4 | 67.2 | 0.405 |
| Sex |  |  |  |  |  |  |  |  |  |
| female | 677 | 47.7 | 359 | 144.3 | 68.2 | 318 | 139.9 | 64.3 | 0.193 |
| male | 742 | 52.3 | 404 | 142.5 | 71.4 | 338 | 144.9 | 69.8 | 0.323 |
| Trauma |  |  |  |  |  |  |  |  |  |
| non-trauma | 892 | 62.9 | 471 | 145.3 | 66.2 | 421 | 147.6 | 66.8 | 0.298 |
| trauma | 527 | 37.1 | 292 | 140.2 | 75.5 | 235 | 133.1 | 67.0 | 0.132 |
| TEP<sup>‡</sup> |  |  |  |  |  |  |  |  |  |
| 0 | 1,052 | 74.1 | 551 | 141.6 | 70.8 | 501 | 141.3 | 65.1 | 0.466 |
| 1 | 308 | 21.7 | 180 | 144.3 | 63.0 | 128 | 146.9 | 73.2 | 0.366 |
| more than 2 | 59 | 4.2 | 32 | 167.1 | 82.0 | 27 | 142.9 | 76.3 | 0.131 |
| hospitalization |  |  |  |  |  |  |  |  |  |
| return home | 710 | 50.0 | 381 | 126.3 | 63.7 | 329 | 123.3 | 65.2 | 0.267 |
| hospitalization | 709 | 50.0 | 382 | 160.3 | 71.6 | 327 | 161.7 | 63.6 | 0.390 |
| Consultation<sup>§</sup> |  |  |  |  |  |  |  |  |  |
| 0 | 340 | 24.0 | 129 | 125.9 | 61.2 | 211 | 132.6 | 55.1 | 0.147 |
| 1 | 806 | 56.8 | 455 | 137.1 | 69.2 | 351 | 138.7 | 69.3 | 0.372 |
| 2 | 225 | 15.9 | 147 | 167.2 | 71.3 | 78 | 174.3 | 74.6 | 0.244 |
| more than 3 | 48 | 3.4 | 32 | 192.0 | 59.7 | 16 | 198.1 | 59.6 | 0.370 |
| XP |  |  |  |  |  |  |  |  |  |
| without | 204 | 14.4 | 105 | 104.3 | 65.7 | 99 | 97.2 | 59.1 | 0.208 |
| with | 1,215 | 85.6 | 658 | 149.5 | 68.5 | 557 | 150.5 | 65.4 | 0.404 |
| CT |  |  |  |  |  |  |  |  |  |
| without | 489 | 34.5 | 252 | 112.7 | 58.5 | 237 | 110.5 | 58.4 | 0.343 |
| with | 930 | 65.5 | 511 | 158.5 | 70.1 | 419 | 160.5 | 56.1 | 0.323 |
| MRI |  |  |  |  |  |  |  |  |  |
| without | 1,292 | 91.1 | 696 | 138.2 | 67.5 | 596 | 136.0 | 61.6 | 0.268 |
| with | 127 | 8.9 | 67 | 196.5 | 72.0 | 60 | 206.8 | 84.5 | 0.230 |
| Blood test |  |  |  |  |  |  |  |  |  |
| without | 162 | 11.4 | 70 | 79.5 | 39.0 | 92 | 84.2 | 51.4 | 0.262 |
| with | 1,257 | 88.6 | 693 | 149.8 | 69.1 | 564 | 152.0 | 64.6 | 0.284 |
| Catheterization |  |  |  |  |  |  |  |  |  |
| without | 1,413 | 99.6 | 760 | 143.6 | 69.9 | 653 | 142.6 | 67.2 | 0.396 |
| with | 6 | 0.4 | 3 | 83.3 | 39.3 | 3 | 110.7 | 51.4 | 0.253 |
| Endoscopy |  |  |  |  |  |  |  |  |  |
| without | 1,391 | 98.0 | 748 | 142.9 | 70.1 | 643 | 142.0 | 66.7 | 0.407 |
| with | 28 | 2.0 | 15 | 166.1 | 53.8 | 13 | 164.5 | 89.0 | 0.476 |

t-test
LOS= length of stay; minutes, n= number, SD= standard deviation, JNP= Japanese nurse practitioner
XP= x-ray photography, CT= computerized tomography, MRI= magnetic resonance imaging
<sup>‡</sup>TEP= tertiary emergency patients; number of tertiary emergency patients when the subjects arrived at the hospital.<sup>§</sup>Consultation= number of consultations with departments other than the emergency department.

### The relationship between the length of stay and care leader

Correlations between the independent variables were examined to avoid multicollinearity in the multiple regression analysis. The variables that had a significant correlation with the length of stay were age, trauma, outcomes, consultation, and XP, CT, MRI, and blood tests. The correlations ranged from −0.066 (p < .05) to 0.331 (p < .01; Table 4).

**Table 4.** Correlation coefficient between variables (N=1,419)

| Variables | LOS | Cl | Age | Sex | Trauma | TEP | Oc | Cons | XP | CT | MRI | Bt | Cath |
| --- | --- | --- | --- | --- | --- | --- | --- | --- | --- | --- | --- | --- | --- |
| Care leader | 0.006 |  |  |  |  |  |  |  |  |  |  |  |  |
| Age | 0.244<sup>b</sup> | 0.013 |  |  |  |  |  |  |  |  |  |  |  |
| Sex | 0.010 | 0.022 | -0.069<sup>b</sup> |  |  |  |  |  |  |  |  |  |  |
| Trauma | -0.066<sup>b</sup> | 0.041 | -0.259<sup>b</sup> | -0.119<sup>b</sup> |  |  |  |  |  |  |  |  |  |
| TEP | 0.033 | 0.032 | -0.021 | 0.000 | -0.034 |  |  |  |  |  |  |  |  |
| Outcomes | 0.262<sup>b</sup> | 0.003 | 0.259<sup>b</sup> | 0.116<sup>a</sup> | -0.192<sup>b</sup> | -0.060<sup>a</sup> |  |  |  |  |  |  |  |
| Consultation | 0.202<sup>b</sup> | 0.176<sup>b</sup> | 0.103<sup>b</sup> | -0.012 | 0.152<sup>b</sup> | -0.004 | -0.018 |  |  |  |  |  |  |
| XP | 0.251<sup>b</sup> | 0.037 | 0.299<sup>b</sup> | 0.021 | -0.010 | -0.045 | **0.754<sup>b</sup>** | 0.092<sup>b</sup> |  |  |  |  |  |
| CT | 0.331<sup>b</sup> | 0.053 | 0.206<sup>b</sup> | 0.085 | -0.078 | -0.056<sup>a</sup> | 0.364<sup>b</sup> | 0.186<sup>b</sup> | 0.223<sup>b</sup> |  |  |  |  |
| MRI | 0.267<sup>b</sup> | -0.014 | 0.898<sup>b</sup> | 0.028 | -0.275<sup>b</sup> | 0.001 | 0.374<sup>b</sup> | 0.070<sup>b</sup> | 0.183<sup>a</sup> | 0.475<sup>b</sup> |  |  |  |
| Blood test | 0.318<sup>b</sup> | 0.156<sup>b</sup> | 0.414<sup>b</sup> | 0.107 | **0.596<sup>b</sup>** | -0.041 | **0.860<sup>b</sup>** | 0.034 | **0.552<sup>b</sup>** | **0.581<sup>b</sup>** | 0.427<sup>b</sup> |  |  |
| Catheterization | -0.044 | -0.032 | 0.027 | **1.000<sup>a</sup>** | **-1.000<sup>a</sup>** | 0.022 | **1.000** | 0.001 | **1.000** | -0.136 | 0.130 | **1.000** |  |
| Endoscopy | 0.046 | -0.002 | 0.043 | 0.126 | **-0.513<sup>b</sup>** | 0.049 | 0.407 | 0.022 | **1.000<sup>a</sup>** | -0.233<sup>a</sup> | -0.179 | **1.000** | **-1.000** |

For the continuous variables, Spearman's correlation coefficient were calculated between them.
The continuous and binary variables were calculated Point-biserial correlations coefficients.
Binary variables and binary variables were calculated Tetrachoric correlation coefficient.
<sup>a</sup>p<.05, <sup>b</sup>p<.01, bold: an absolute value of 0.500 or more
N= number, LOS= length of stay;minutes, Cl= Care leader, Oc= Outcomes, Cons= Consultation, Bt= Blood test, Cath= Catheterizaion
TEP= tertiary emergency patients; the number of tertiary emergency patients when the subjects arrived at the hospital.
XP= x-ray photography, CT= computerized tomography, MRI= magnetic resonance imaging
The reference of care leader variable was "physician trainees".
The reference of sex variable was "female".

To clarify the relationship between the length of stay and each variable, a simple linear regression analysis was performed using the length of stay as the dependent variable. No significant relationship was found between the length of stay and care leader (unstandardized coefficient: 0.88; 95% CI: −6.29 to 8.05). The variables with significant associations were age, consultation, MRI, blood test, trauma, outcomes, CT, and XP (Table 5).

**Table 5.** Association of length of stay in the ED with care provider (N=1,419)

| Variables |  | n | crude | adjusted |  |
| --- | --- | --- | --- | --- | --- |
|  |  |  |  | Model 1 | Model 2 |
|  |  |  | B (95%CI) | B(95%CI) | B(95%CI) |
| Care leader | physicians trainees | 656 | reference | reference | reference |
|  | JNP | 763 | 0.88(-6.29—8.05) | -6.67(-13.21—-0.13) | -6.76(-13.30—-0.22) |
| Age | year |  | 0.66(0.51—0.81) | 0.23(0.08—0.38) | 0.25(0.10—0.41) |
| Sex | female | 677 | reference | reference | reference |
|  | male | 742 | 1.33(-5.82—8.49) | 0.08(-6.36—6.53) | 0.51(-5.95—6.97) |
| TEP | person |  | 4.16(-2.18—10.49) | 6.30(0.62—11.99) | 6.54(0.85—12.23) |
| Endoscopy | without | 1391 | reference | reference | reference |
|  | with | 28 | 22.89(-2.78—48.56) | 14.23(-8.86—37.31) | 16.05(-7.11—39.21) |
| Consultation | case |  | 19.80(15.11—24.49) | 17.60(13.20—22.00) | 16.84(12.36—21.32) |
| MRI | without | 1292 | reference | reference | reference |
|  | with | 127 | 64.18(52.11—76.25) | 53.26(41.96—64.56) | 54.23(42.88—65.58) |
| Blood test | without | 162 | reference | reference | reference |
|  | with | 1257 | 68.62(57.96—79.27) | 56.47(45.31—67.64) | 58.73(47.29—70.17) |
| Trauma | without | 892 | reference |  | reference |
|  | with | 527 | -9.37(-16.75—-1.99) |  | 6.44(-0.82—13.70) |
| Outcomes | return home | 710 | reference |  |  |
|  | hospitalization | 709 | 35.97(29.07—42.87) |  |  |
| CT | without | 489 | reference |  |  |
|  | with | 930 | 47.77(40.67—54.86) |  |  |
| XP | without | 204 | reference |  |  |
|  | with | 1215 | 49.10(39.24—58.97) |  |  |
| Catheterization | without | 1413 | reference |  |  |
|  | with | 6 | -46.11(-101.15—8.92) |  |  |

crude: simple linear regression analysis
adjusted: multiple linear regression analysis; the models that did not include all variables that were correlated above 0.5 at the same time.
Model1: $R^2 = 0.20$ , VIF= 1.01~1.24, Model2: $R^2 = 0.21$ , VIF= 1.01~1.30
dependent variable= Length of stay (minutes), ED= emergency department, N and n= number
B= unstandardized regression coefficient, CI= confidence interval for B, JNP= Japanese nurse practitioner
TEP= tertiary emergency patients; number of tertiary emergency patients when the subject arrives at the hospital
Consultation= number of consultations with departments other than the emergency department, MRI= magnetic resonance imaging
CT= computerized tomography, XP= x-ray photography
S1 Table, we showed the results of multiple linear regression analysis for exploring confounding factors for LOS and care providers.

A multiple linear regression analysis was performed to clarify the relationship between the length of stay and care leader after adjusting for other variables. To avoid multicollinearity in the multiple regression analysis, we built the models so that no variables with absolute values of correlation coefficients greater than 0.500 were included concurrently. The length of stay was the dependent variable, and the independent variables were care leader, age, sex, tertiary emergency patients, endoscopy, consultation, MRI, and blood test.

The results of the multiple linear regression analysis demonstrated that there was a significant relationship between the length of stay and the care leader. The length of stay of the patients was significantly shorter by six minutes in the JNP group compared to the physician trainees’ group (−6.67; −13.21 to −0.13). The other significant variables were age (0.23; 0.08 to 0.38), TEPs (6.30; 0.62 to 11.99), consultation (17.60; 13.20 to 22.00), MRI (53.26; 41.96 to 64.56), and blood test (56.47; 45.31 to 67.64; Table 5).

### Relationship between care leaders and consultation, and Robustness of the sign of the care leader’s coefficients

Due to the sign reversal in the crude and adjusted models for care leader, a multiple linear regression analysis was conducted to explore variables that have a relationship with care leader regarding the length of stay. The sign of the care leader changed to negative only in the model with the inclusion of a consultation (S1 Table). We constructed Model 2 with the addition of trauma; however, the sign of the care leader coefficients did not change. Additionally, we demonstrated that, in Model 2, replacing trauma with outcomes, CT, or the XP variables did not change the sign of the care leaders’ coefficients (S2 Table).

## Discussion

This study demonstrated that patients’ length of stay was significantly shorter by six minutes in the JNP group than in the physician trainees group. In any of the multiple regression models that trauma, outcomes, CT, and XP were entered into, the sign of care leaders’ coefficient remained the same and was negative. This suggests that the efficiency of JNP-led care is more efficient than the care led by physician trainees. To our knowledge, this was the first study to demonstrate the outcome of JNP-led care in a secondary ED in Japan. However, it should be noted that all patients received care at a secondary ED during weekday shifts.

The allegation in the following studies was that the NPs had a beneficial effect on patients’ length of stay. A study conducted in the ED of a small city found that the median length of stay of the NP-provided patient group was 77 minutes, compared to the 174 minutes of the NP and emergency physician-provided group [12]. Further, a cohort study in the ED of a community hospital in an urban area revealed the following findings: 85 ± 56 minutes in the physician group and 65 ± 42 in the emergency NP group [13]. A study of outpatients in the ED with levels ranging from non-urgent to potentially life-threatening revealed a median length of stay of 94 minutes in the NP-provided care group and 170 minutes in the patient group who received conventional care [14]. Although the results cannot be fully generalized because our study was conducted in Japan and NPs’ work styles and work contents may differ, our findings support previous studies that NP-provided care is associated with shortened length of stay among ED patients.

Concerning the method, some previous studies adjusted the bias of patients’ severity and urgency by stratifying the NP-provided medical care group and physicians’ medical care group by triage category [12,14,15]. Unlike previous studies, to address these biases, the current study limited participants to patients who were treated at a secondary ED. In addition, we adjusted for patients’ characteristics and whether they received tests in the multivariate analyses, which still revealed that the NP-led group had a significantly shorter length of stay than the physician trainees-led group.

The following previous studies also suggest that NP-provided care is not inferior to physician-provided care. A case-control study of 725 emergency outpatients who came home with orthopedic disease demonstrated that the median length of stay in the conventional system without NP was 137 minutes and 125 minutes with NP, which was non-significant [24]. Steiner et al. [12] found that the length of stay was 123 minutes for the emergency physician-provided patients and 125 minutes for those provided with care by the emergency physicians and NP, which was non-significant (p = .13). In contrast, in a study investigating patients who returned home after visiting the ambulatory care clinic, the median length of stay for the physician-provided group was 143 minutes, and the NP-provided was 156 minutes, which was a significant difference [15]. In one study that evaluated physicians’ and NPs’ care for minorly injured patients in an emergency facility, the average care time of the physician group was 12 minutes shorter than the NP group [25]. In these previous studies, the NP-provided group had a longer length of stay than the physician-provided group in the ED, which contrasts our findings. We cannot deny the existence of publication bias—that only favorable results concerning NPs are published. However, our study suggests that there is no disadvantage to patients’ length of stay by receiving care from a JNP instead of a physician.

The length of stay is defined by the performance of four processes: medical care in the ED, medical care of specialized medical departments, ward coordination, and movement. In particular, the medical care in the ED includes the time related to interviews, physical examinations, judgment and implementation of various tests, interpretation of test results, provisional diagnosis, consultation with other departments, and treatment policy decisions.

Therefore, the length of stay reflects not only the power of the department in charge but also the speed of cooperation among various departments within the hospital.

The results of our study, adjusted for patients’ characteristics and whether medical tests were conducted, suggested that the care led by the JNP could shorten patients’ length of stay. As illustrated in S1 Table, the coefficients’ direction was negative when the independent variables were care leader and consultation variables. The JNP group tended to spend about four minutes less time in the ED than the physician trainee group concerning consultations with other departments. The shorter length of stay may be owing to the relationship between other departments and other factors depending on the skill and experience of the individual JNP and their ability to coordinate. This is probably because JNPs collaborate with various departments in the hospital more accurately and quickly than physician trainees. The JNPs are more likely to have more years of employment in hospitals than physician trainees, and they may be familiar with the hospital system and culture. The results suggest that JNPs are equivalent or greater to physician trainees in their efficiency and ability to manage patients.

A previous study compared NPs and nurses and examined patients’ length of stay in the ED [38]. In that study, the mean length of stay of patients with NP-led care was 73 minutes, while that of patients with nurse-led care was 98 minutes. The NPs could assess the test order, interpret test results, and provide medical care (including triage). Certainly, nurses may reduce patients’ length of stay in EDs by improving manuals, operations, and triage skills. However, NPs have a better ability to understand pathological conditions, provide necessary tests, and provide predictive care than nurses, and they are often involved in medical care. Therefore, it was considered that they are directly involved in decreasing patients’ length of stay.

Another advantage of introducing JNPs in the ED is that the quality of medical care is improved. For example, NPs can lead medical care for patients with mild conditions, and physicians can focus on treating patients with more severe and complicated diseases. In recent years, there has been a shortage of emergency physicians owing to the uneven distribution of physicians [39], and a work style reform law has promoted restrictions on physicians’ ability to work overtime [40]. Consequently, JNPs reduce emergency physicians’ work-related burdens. In addition, JNPs enable smoother collaboration and task sharing between physicians and nurses. The introduction of JNPs will lead to investment in hospital organizations and benefit patients, families, and team care [41]. Thus, policymakers should consider introducing JNP in secondary emergency outpatients to address the limitations mentioned above in medical settings.

### The relationship between the length of stay and other factors

Age increased by 0.2 minutes with an increase of 1 year. An increase in the number of TEPs increased the length of stay by about six minutes. Since the two emergency nurses were concurrently working in the tertiary and secondary ED, this extension of time was thought to be related to the lack of manpower. In addition, TEPs are given priority over secondary-emergency patients for imaging examinations and other tests, which may have increased the waiting time for secondary-emergency patients, thereby prolonging their length of stay. The number of consultations with other departments increased by approximately 17 minutes, suggesting that the length of stay is extended by the time it takes to be examined by a specialist in another department. The length of stay in the ED was about 53 minutes longer for those who had MRI than for those who did not, and about 56 minutes longer for those who had blood tests than for those who did not. It suggested that the MRI themselves took time and the blood tests took time to determine the results, which prolonged the length of stay in the ED.

### Strengths

The strengths of this study are the number of participants and the use of objective medical record data. We assessed one JNP at one target facility; therefore, the internal validity is maintained. In addition, we can compare our results to those of previous studies conducted overseas.

## Limitations

As this was a retrospective survey, the accuracy and completeness of the data are lacking owing to the difference in record keeping. Further, it is undeniable that the data may comprise typing errors as it was extracted from the emergency outpatient transport patient records and entered into the research dataset. Moreover, the external generalizability of our results is limited since our data reflect the practice of one JNP working at one site. Patients were admitted during the day on weekdays; therefore, data concerning nighttime and weekend/holiday cases were not included. The results should thus be generalized with caution. Further, a multivariate analysis was performed using nine variables. The coefficient of determination (*R*^*2*^) of the multiple linear regression analysis model in this study was 0.20. The remaining 80% of the length of stay was explained by other factors that were not surveyed in this study. This study focused only on outpatient time, and we did not consider other important outcomes. In addition, we were unable to investigate the background of the physician trainees, including their years of experience.

## Conclusion

The length of stay of patients was significantly shorter by six minutes in the JNP group compared to the physician trainees’ group. This time difference suggests that the medical care led by JNP is more efficient than that led by physician trainees. In the future, the cost-effectiveness of JNP’s medical care (e.g., salary and remuneration obtained from medical care), safety (e.g., consistency between initial diagnosis and post-hospitalization diagnosis and return-to-home rate of returning patients), and patient satisfaction should be examined in a multi-institutional joint study.

## Data Availability

The data that support the findings of this study are available from the corresponding author [KU] upon reasonable request.

## Acknowledgments

Our special thanks to the patients who participated in the study. Special thanks are also extended to the staff members of the Department of Health Sciences, University of Yamanashi, for their any suggestions and advices to brush up on this study. We also thank Editage(www.editage.com) for English language editing.

## Supporting information

S1 Table. Association of length of stay in the ED with care provider and each variable.

S2 Table. Association of length of stay in the ED with care leader, replacing trauma with either Outcomes, CT, or XP variables.

**S1 Table.**
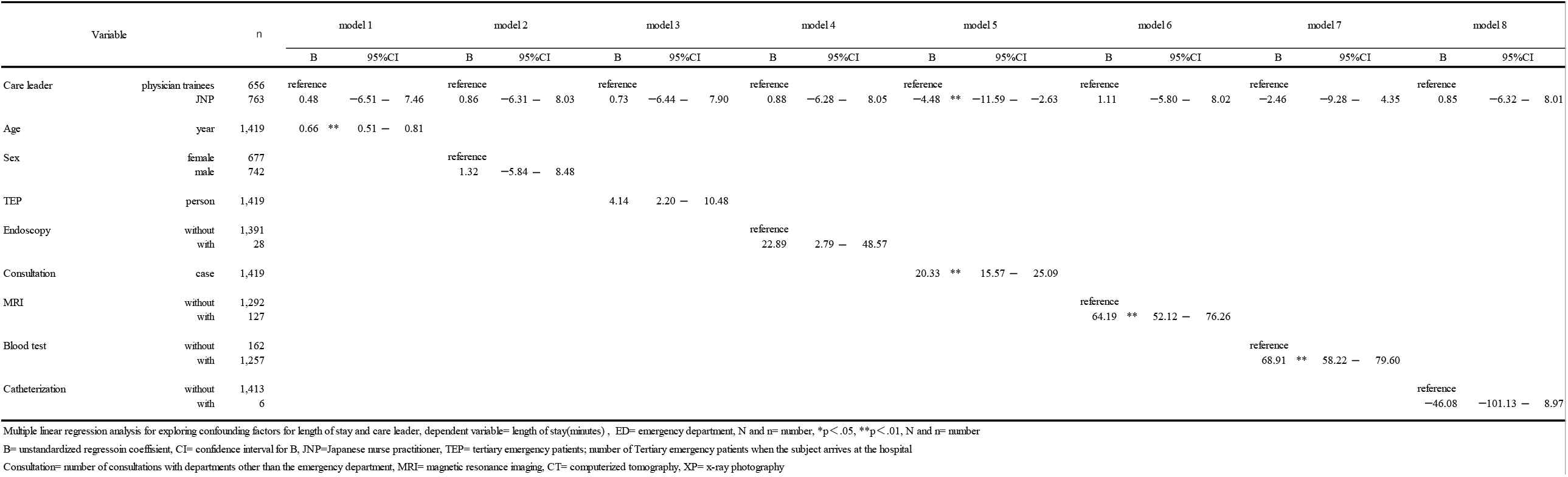
Association of length of stay in the ED with care leader and each variable (N=1,419)

**S2 Table.**
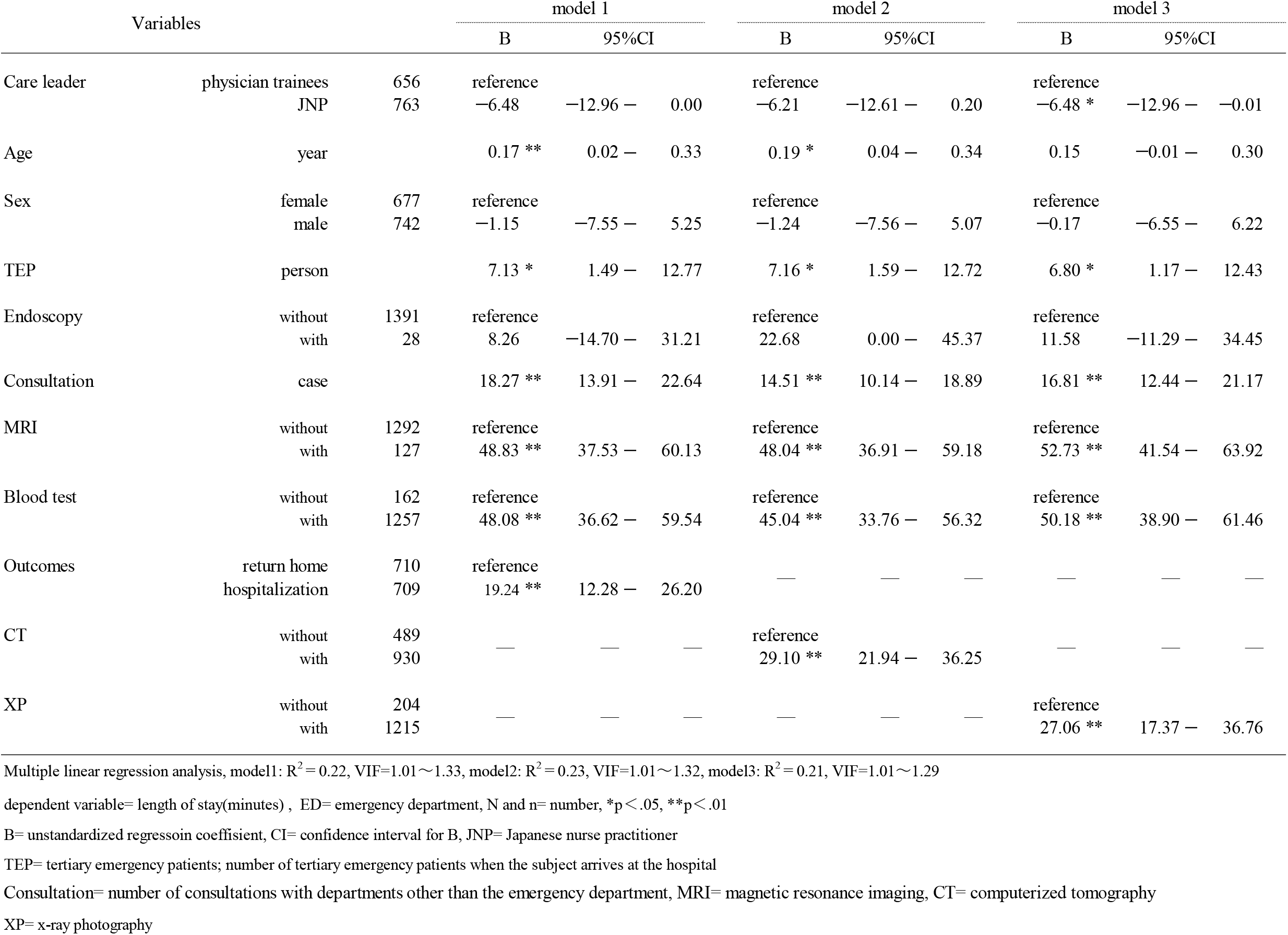
Association of length of stay in the ED with care leader, replacing trauma with either Outcomes, CT, or XP variables (N=1,419)

| Variables |  |  | model 1 |  |  | model 2 |  |  | model 3 |  |  |
| --- | --- | --- | --- | --- | --- | --- | --- | --- | --- | --- | --- |
|  |  |  | B | 95% CI |  | B | 95% CI |  | B | 95% CI |  |
| Care leader | physician trainees | 656 | reference |  |  | reference |  |  | reference |  |  |
|  | JNP | 763 | −6.48 | −12.96 | 0.00 | −6.21 | −12.61 | 0.20 | −6.48 * | −12.96 | −0.01 |
| Age | year |  | 0.17 ** | 0.02 | 0.33 | 0.19 * | 0.04 | 0.34 | 0.15 | −0.01 | 0.30 |
| Sex | female | 677 | reference |  |  | reference |  |  | reference |  |  |
|  | male | 742 | −1.15 | −7.55 | 5.25 | −1.24 | −7.56 | 5.07 | −0.17 | −6.55 | 6.22 |
| TEP | person |  | 7.13 * | 1.49 | 12.77 | 7.16 * | 1.59 | 12.72 | 6.80 * | 1.17 | 12.43 |
| Endoscopy | without | 1391 | reference |  |  | reference |  |  | reference |  |  |
|  | with | 28 | 8.26 | −14.70 | 31.21 | 22.68 | 0.00 | 45.37 | 11.58 | −11.29 | 34.45 |
| Consultation | case |  | 18.27 ** | 13.91 | 22.64 | 14.51 ** | 10.14 | 18.89 | 16.81 ** | 12.44 | 21.17 |
| MRI | without | 1292 | reference |  |  | reference |  |  | reference |  |  |
|  | with | 127 | 48.83 ** | 37.53 | 60.13 | 48.04 ** | 36.91 | 59.18 | 52.73 ** | 41.54 | 63.92 |
| Blood test | without | 162 | reference |  |  | reference |  |  | reference |  |  |
|  | with | 1257 | 48.08 ** | 36.62 | 59.54 | 45.04 ** | 33.76 | 56.32 | 50.18 ** | 38.90 | 61.46 |
| Outcomes | return home | 710 | reference |  |  | — | — | — | — | — | — |
|  | hospitalization | 709 | 19.24 ** | 12.28 | 26.20 | — | — | — | — | — | — |
| CT | without | 489 | — | — | — | reference |  |  | — | — | — |
|  | with | 930 | — | — | — | 29.10 ** | 21.94 | 36.25 | — | — | — |
| XP | without | 204 | — | — | — | — | — | — | reference |  |  |
|  | with | 1215 | — | — | — | — | — | — | 27.06 ** | 17.37 | 36.76 |

Multiple linear regression analysis, model1: $R^2 = 0.22$ , VIF=1.01~1.33, model2: $R^2 = 0.23$ , VIF=1.01~1.32, model3: $R^2 = 0.21$ , VIF=1.01~1.29
dependent variable= length of stay(minutes) , ED= emergency department, N and n= number, \* $p < .05$ , \*\* $p < .01$
B= unstandardized regressoin coefficient, CI= confidence interval for B, JNP= Japanese nurse practitioner
TEP= tertiary emergency patients; number of tertiary emergency patients when the subject arrives at the hospital
Consultation= number of consultations with departments other than the emergency department, MRI= magnetic resonance imaging, CT= computerized tomography
XP= x-ray photography

